## Supplementary data for "A machine-learning method for biobank-scale genetic prediction of blood group antigens"

#### SUPPLEMENTARY MATERIALS

Hyvärinen K[1*], Haimila K[2], Moslemi C[3,4], Blood Service Biobank[5], Olsson ML[6,7], Ostrowski SR[8,9], Pedersen OB[3,9], Erikstrup C[10], Partanen J[1], Ritari J[1]

1. *Research and Development, Finnish Red Cross Blood Service, Helsinki, Finland*
2. *Blood Group Unit, Finnish Red Cross Blood Service, Vantaa, Finland*
3. *Department of Clinical Immunology, Zealand University Hospital,* Køge, *Denmark*
4. *Department of Clinical Immunology, Aarhus University Hospital, Aarhus, Denmark*
5. *Finnish Red Cross Blood Service, Vantaa, Finland*
6. *Department of Laboratory Medicine, Lund University, Lund, Sweden*
7. *Department of Clinical Immunology and Transfusion Medicine, Office for Medical Services, Region Skåne, Sweden*
8. *Department of Clinical Immunology, Copenhagen University Hospital, Rigshopitalet, Copenhagen, Denmark*
9. *Department of Clinical Medicine, University of Copenhagen, Copenhagen, Denmark*
10. *Department of Clinical Immunology, Aarhus University Hospital, Skejby, Denmark*

Short title: Blood group prediction method

*Corresponding author:
Kati Hyvärinen, PhD, Associate professor
Finnish Red Cross Blood Service
Research and Development
Biomedicum Helsinki 1, Haartmaninkatu 8, 00290 Helsinki, FINLAND


Table of contents 2 ........ **Error! Bookmark not defined.**

### Supplementary Table 1. Blood group/HPA-1 genes and genetic regions

| Blood group/HPA system | Ensembl gene ID | Genes analyzed | CHR | Start position -2 Kbp GRCh38 | End position +2 Kbp GRCh38 |
|---|---|---|---|---|---|
| ABO | ENSG00000175164 | *ABO* | 9 | 133231278 | 133278024 |
| CartwrightYt | ENSG00000087085 | *ACHE* | 7 | 100887994 | 100898974 |
| Colton | ENSG00000240583 | *AQP1* | 7 | 30909853 | 30927517 |
| Diego | ENSG00000004939 | *SLC4A1* | 17 | 44246390 | 4427014 |
| Dombrock | ENSG00000111339 | *ART4* | 12 | 14823569 | 14845526 |
| Duffy | ENSG00000213088 | *ACKR1* | 1 | 159201307 | 159208500 |
| Gerbich | ENSG00000136732 | *GYPC* | 2 | 126654133 | 126698667 |
| HPA-1 | ENSG00000259207 | *ITGB3* | 17 | 47251827 | 47315743 |
| Kell | ENSG00000197993 | *KEL* | 7 | 142939114 | 142964363 |
| Knops | ENSG00000203710 | *CR1* | 1 | 207494128 | 207643765 |
| Kidd | ENSG00000141469 | *SLC14A1* | 18 | 45685025 | 45754520 |
| Landsteiner-Wiener | ENSG00000105371 | *ICAM4* | 19 | 10284955 | 10290522 |
| Lewis | ENSG00000176920 | *FUT2* | 19 | 48693971 | 48707951 |
| Lewis | ENSG00000171124 | *FUT3* | 19 | 5840888 | 5853474 |
| Lutheran | ENSG00000187244 | *BCAM* | 19 | 44807071 | 44823421 |
| MNS | ENSG00000170180 | *GYPA* | 4 | 144107303 | 144142751 |
| MNS | ENSG00000250361 | *GYPB* | 4 | 143994104 | 144021380 |
| MNS | ENSG00000197465 | *GYPE* | 4 | 143868864 | 143907563 |
| P1PK | ENSG00000128274 | *A4GALT* | 22 | 42690121 | 42723298 |
| P1PK | ENSG00000169255 | *B3GALNT1* | 3 | 161081883 | 161107411 |
| Rh | ENSG00000188672 | *RHCE* | 1 | 25360249 | 25432192 |
| Rh | ENSG00000187010 | *RHD* | 1 | 25270393 | 25332445 |
| VEL | ENSG00000235169 | *SMIM1* | 1 | 3770749 | 3777982 |

CHR, chromosome; GRCh38, Genome Reference Consortium Human Build 38; HPA, human platelet antigen

### Supplementary Table 2. Characteristics of the Danish classification models.

| Blood group/HPA system | Antigen[a] | Genes analyzed | n(variants available) | n(model variants) | Prediction error[b] |
|---|---|---|---|---|---|
| ABO | ABO | *ABO* | 615 | 603 | 1.80E-03 |
| ABO | $A_1$ | *ABO* | 615 | 586 | 1.17E-02 |
| ABO | $A_2$ | *ABO* | 615 | 480 | 5.95E-02 |
| Cartwright | $Yt^a$ | *ACHE* | 76 | 21 | 1.47E-04 |
| Cartwright | $Yt^b$ | *ACHE* | 76 | 44 | 6.15E-04 |
| Colton | $Co^a$ | *AQP1* | 69 | 36 | 9.20E-04 |
| Colton | $Co^b$ | *AQP1* | 69 | 41 | 8.33E-03 |
| Dombrock | $Do^a$ | *ART4* | 141 | 104 | 8.60E-04 |
| Dombrock | $Do^b$ | *ART4* | 141 | 113 | 3.79E-04 |
| Duffy | $Fy^a$ | *ACKR1* | 42 | 42 | 2.42E-03 |
| Duffy | $Fy^b$ | *ACKR1* | 42 | 40 | 7.76E-03 |
| HPA-1 | HPA-1a | *ITGB3* | 377 | 20 | 2.06E-04 |
| HPA-1 | HPA-1b | *ITGB3* | 377 | 91 | 2.28E-03 |
| Kell | K | *KEL* | 87 | 86 | 8.50E-03 |
| Kell | $Kp^a$ | *KEL* | 87 | 50 | 2.75E-03 |
| Kidd | $Jk^a$ | *SLC14A1* | 509 | 504 | 2.47E-03 |
| Kidd | $Jk^b$ | *SLC14A1* | 509 | 492 | 2.09E-03 |
| Knops | $Kn^a$ | *CR1* | 622 | 110 | 4.28E-04 |
| Knops | $Kn^b$ | *CR1* | 622 | 31 | 2.16E-05 |
| Lewis | $Le^a$ | *FUT2. FUT3* | 210 | 206 | 6.48E-03 |
| Lewis | $Le^b$ | *FUT2. FUT3* | 210 | 203 | 1.68E-01 |
| Lutheran | $Lu^a$ | *BCAM* | 98 | 95 | 7.89E-03 |
| Lutheran | $Lu^b$ | *BCAM* | 98 | 77 | 1.49E-03 |
| MNS | M | *GYPA. GYPB. GYPC* | 766 | 736 | 3.58E-03 |
| MNS | N | *GYPA. GYPB. GYPC* | 766 | 743 | 1.15E-02 |
| MNS | S | *GYPA. GYPB. GYPC* | 766 | 702 | 2.54E-03 |
| MNS | s | *GYPA. GYPB. GYPC* | 766 | 655 | 1.72E-03 |
| P1PK | P1 | *A4GALT. B3GALNT1* | 499 | 489 | 1.89E-02 |
| Rh | C | *RHCE. RHD* | 465 | 452 | 2.14E-03 |
| Rh | $C^w$ | *RHCE. RHD* | 465 | 411 | 3.50E-03 |
| Rh | D | *RHCE. RHD* | 465 | 461 | 1.66E-03 |
| Rh | E | *RHCE. RHD* | 465 | 464 | 2.70E-03 |
| Rh | e | *RHCE. RHD* | 465 | 423 | 9.04E-04 |
| Vel | Vel | *SMIM1* | 78 | 49 | 9.48E-04 |

[a] O, $A_1$, and $A_2$ in this column refer to phenotype.

[b] Misclassification frequency obtained from out-of-bag data.

#### Supplementary Table 3. Accuracy metrics for the Finnish models in the Finnish train data set.

| Blood group/HPA system | Antigen[a] | Sensitivity | Specificity | Positive predictive value | Negative predictive value | Balanced accuracy |
|---|---|---|---|---|---|---|
| ABO | A | 1.000 | 1.000 | 1.000 | 1.000 | 1.000 |
| ABO | $A_1$ | 1.000 | 1.000 | 1.000 | 1.000 | 1.000 |
| ABO | $A_2$ | 1.000 | 1.000 | 1.000 | 1.000 | 1.000 |
| ABO | AB | 1.000 | 1.000 | 1.000 | 1.000 | 1.000 |
| ABO | B | 1.000 | 0.955 | 0.929 | 1.000 | 0.977 |
| ABO | O | 1.000 | 0.929 | 0.957 | 1.000 | 0.964 |
| Cartwright | $Yt^b$ | 1.000 | 1.000 | 1.000 | 1.000 | 1.000 |
| Colton | $Co^a$ | 0.000 | 1.000 | NA | 0.997 | 0.500 |
| Colton | $Co^b$ | 1.000 | 1.000 | 1.000 | 1.000 | 1.000 |
| Dombrock | $Do^a$ | 1.000 | 1.000 | 1.000 | 1.000 | 1.000 |
| Dombrock | Dob | 1.000 | 1.000 | 1.000 | 1.000 | 1.000 |
| Duffy | $Fy^a$ | 1.000 | 1.000 | 1.000 | 1.000 | 1.000 |
| Duffy | $Fy^b$ | 0.986 | 0.998 | 0.993 | 0.995 | 0.992 |
| Gerbich | $Ls^a$ | 1.000 | 0.667 | 0.989 | 1.000 | 0.833 |
| HPA-1 | HPA-1a | 1.000 | 1.000 | 1.000 | 1.000 | 1.000 |
| HPA-1 | HPA-1b | 0.944 | 1.000 | 1.000 | 0.933 | 0.972 |
| Kell | K | 1.000 | 1.000 | 1.000 | 1.000 | 1.000 |
| Kell | $Kp^a$ | 1.000 | 1.000 | 1.000 | 1.000 | 1.000 |
| Kell | $Ul^a$ | 1.000 | 1.000 | 1.000 | 1.000 | 1.000 |
| Kidd | $Jk^a$ | 1.000 | 1.000 | 1.000 | 1.000 | 1.000 |
| Kidd | $Jk^b$ | 1.000 | 1.000 | 1.000 | 1.000 | 1.000 |
| Landsteiner-Wiener | $LW^b$ | 1.000 | 1.000 | 1.000 | 1.000 | 1.000 |
| Lewis | $Le^a$ | 1.000 | 1.000 | 1.000 | 1.000 | 1.000 |
| Lewis | $Le^b$ | 0.920 | 0.990 | 0.958 | 0.981 | 0.955 |
| Lutheran | $Lu^a$ | 1.000 | 0.952 | 0.998 | 1.000 | 0.976 |
| MNS | M | 0.972 | 0.998 | 0.986 | 0.996 | 0.985 |
| MNS | N | 0.983 | 0.994 | 0.992 | 0.989 | 0.989 |
| MNS | S | 1.000 | 1.000 | 1.000 | 1.000 | 1.000 |
| MNS | s | 1.000 | 1.000 | 1.000 | 1.000 | 1.000 |
| P1PK | P1 | 0.968 | 0.990 | 0.968 | 0.990 | 0.979 |
| Rh | C | 0.992 | 0.998 | 0.992 | 0.998 | 0.995 |
| Rh | c | 0.982 | 0.994 | 0.993 | 0.985 | 0.988 |
| Rh | $C^w$ | 0.982 | 0.998 | 0.994 | 0.993 | 0.990 |
| Rh | $C^x$ | 1.000 | 1.000 | 1.000 | 1.000 | 1.000 |
| Rh | D | 0.957 | 1.000 | 1.000 | 0.998 | 0.978 |
| Rh | E | 1.000 | 1.000 | 1.000 | 1.000 | 1.000 |
| Rh | e | 0.998 | 1.000 | 1.000 | 0.950 | 0.999 |
| Rh | $hr^B$ | 1.000 | 0.993 | 0.800 | 1.000 | 0.996 |
| Rh | $hr^S$ | 0.952 | 1.000 | 1.000 | 0.998 | 0.976 |

[a] O, $A_1$, and $A_2$ in this column refer to phenotype.

#### Supplementary Table 4. Accuracy metrics for the Finnish models in the Finnish full data set.

| Blood group/HPA system | Antigen[a] | Sensitivity | Specificity | Positive predictive value | Negative predictive value | Balanced accuracy |
|---|---|---|---|---|---|---|
| ABO | A | 0.999 | 1.000 | 1.000 | 0.998 | 0.999 |
| ABO | $A_1$ | 1.000 | 1.000 | 1.000 | 1.000 | 1.000 |
| ABO | $A_2$ | 1.000 | 1.000 | 1.000 | 1.000 | 1.000 |
| ABO | AB | 1.000 | 0.998 | 0.999 | 1.000 | 0.999 |
| ABO | B | 1.000 | 1.000 | 1.000 | 1.000 | 1.000 |
| ABO | O | 1.000 | 0.963 | 0.977 | 1.000 | 0.981 |
| Cartwright | $Yt^b$ | 1.000 | 1.000 | 1.000 | 1.000 | 1.000 |
| Colton | $Co^a$ | 1.000 | 1.000 | 1.000 | 1.000 | 1.000 |
| Colton | $Co^b$ | 1.000 | 0.976 | 0.998 | 1.000 | 0.988 |
| Dombrock | $Do^a$ | 1.000 | 1.000 | 1.000 | 1.000 | 1.000 |
| Dombrock | Dob | 1.000 | 1.000 | 1.000 | 1.000 | 1.000 |
| Duffy | $Fy^a$ | 1.000 | 1.000 | 1.000 | 1.000 | 1.000 |
| Duffy | $Fy^b$ | 0.986 | 0.999 | 0.997 | 0.995 | 0.993 |
| Gerbich | $Ls^a$ | 1.000 | 0.833 | 0.995 | 1.000 | 0.917 |
| HPA-1 | HPA-1a | 1.000 | 1.000 | 1.000 | 1.000 | 1.000 |
| HPA-1 | HPA-1b | 0.971 | 1.000 | 1.000 | 0.964 | 0.986 |
| Kell | K | 1.000 | 1.000 | 1.000 | 1.000 | 1.000 |
| Kell | $Kp^a$ | 1.000 | 1.000 | 1.000 | 1.000 | 1.000 |
| Kell | $Ul^a$ | 1.000 | 1.000 | 1.000 | 1.000 | 1.000 |
| Kidd | $Jk^a$ | 1.000 | 1.000 | 1.000 | 1.000 | 1.000 |
| Kidd | $Jk^b$ | 0.997 | 1.000 | 1.000 | 0.999 | 0.999 |
| Landsteiner-Wiener | $LW^b$ | 0.997 | 1.000 | 1.000 | 0.944 | 0.998 |
| Lewis | $Le^a$ | 0.996 | 1.000 | 1.000 | 0.963 | 0.998 |
| Lewis | $Le^b$ | 0.940 | 0.995 | 0.979 | 0.985 | 0.968 |
| Lutheran | $Lu^a$ | 1.000 | 0.976 | 0.999 | 1.000 | 0.988 |
| MNS | M | 0.972 | 0.996 | 0.972 | 0.996 | 0.984 |
| MNS | N | 0.988 | 0.988 | 0.983 | 0.991 | 0.988 |
| MNS | S | 1.000 | 1.000 | 1.000 | 1.000 | 1.000 |
| MNS | s | 1.000 | 1.000 | 1.000 | 1.000 | 1.000 |
| P1PK | P1 | 0.984 | 0.984 | 0.952 | 0.995 | 0.984 |
| Rh | C | 0.996 | 0.998 | 0.992 | 0.999 | 0.997 |
| Rh | c | 0.983 | 0.995 | 0.994 | 0.986 | 0.989 |
| Rh | $C^w$ | 0.976 | 0.999 | 0.997 | 0.991 | 0.988 |
| Rh | $C^x$ | 1.000 | 0.996 | 0.999 | 1.000 | 0.998 |
| Rh | D | 0.978 | 0.999 | 0.978 | 0.999 | 0.988 |
| Rh | E | 0.993 | 1.000 | 1.000 | 0.953 | 0.997 |
| Rh | e | 0.997 | 1.000 | 1.000 | 0.927 | 0.999 |
| Rh | $hr^B$ | 0.871 | 0.995 | 0.818 | 0.996 | 0.933 |
| Rh | $hr^S$ | 0.976 | 0.999 | 0.976 | 0.999 | 0.988 |

[a] O, $A_1$, and $A_2$ in this column refer to phenotype.

#### Supplementary Table 5. Accuracy metrics for the Finnish models in the Danish full data set.

| Blood group/HPA system | Antigen[a] | Sensitivity | Specificity | Positive predictive value | Negative predictive value | Balanced accuracy |
|---|---|---|---|---|---|---|
| ABO | A | 0.993 | 0.998 | 0.998 | 0.991 | 0.995 |
| ABO | $A_1$ | 0.972 | 0.988 | 0.963 | 0.991 | 0.980 |
| ABO | $A_2$ | 0.984 | 0.803 | 0.931 | 0.949 | 0.894 |
| ABO | AB | 0.999 | 0.997 | 1.000 | 0.985 | 0.998 |
| ABO | B | 1.000 | 0.975 | 0.997 | 0.998 | 0.987 |
| ABO | O | 0.998 | 0.992 | 0.993 | 0.998 | 0.995 |
| Cartwright | $Yt^b$ | 0.999 | 0.996 | 1.000 | 0.991 | 0.998 |
| Colton | $Co^a$ | 0.667 | 1.000 | 0.800 | 0.999 | 0.833 |
| Colton | $Co^b$ | 0.999 | 0.854 | 0.985 | 0.989 | 0.926 |
| Dombrock | $Do^a$ | 0.999 | 0.999 | 0.999 | 0.999 | 0.999 |
| Dombrock | $Do^b$ | 0.997 | 1.000 | 0.999 | 1.000 | 0.999 |
| Duffy | $Fy^a$ | 0.963 | 0.996 | 0.992 | 0.982 | 0.979 |
| Duffy | $Fy^b$ | 0.945 | 0.998 | 0.990 | 0.987 | 0.971 |
| HPA-1 | HPA-1a | 1.000 | 1.000 | 1.000 | 1.000 | 1.000 |
| HPA-1 | HPA-1b | 0.997 | 0.881 | 0.950 | 0.993 | 0.939 |
| Kell | K | 0.998 | 0.834 | 0.986 | 0.979 | 0.916 |
| Kell | $Kp^a$ | 1.000 | 0.870 | 0.997 | 0.981 | 0.935 |
| Kidd | $Jk^a$ | 0.991 | 0.998 | 0.995 | 0.997 | 0.995 |
| Kidd | $Jk^b$ | 0.999 | 0.325 | 0.357 | 0.999 | 0.662 |
| Landsteiner-Wiener | $Lw^b$ | 0.000 | 1.000 | NA | 0.010 | 0.500 |
| Lewis | $Le^a$ | 0.981 | 0.982 | 0.996 | 0.914 | 0.981 |
| Lewis | $Le^b$ | 0.465 | 0.994 | 0.983 | 0.707 | 0.729 |
| Lutheran | $Lu^a$ | 0.998 | 0.914 | 0.992 | 0.975 | 0.956 |
| MNS | M | 0.942 | 0.999 | 0.996 | 0.984 | 0.970 |
| MNS | N | 0.966 | 0.928 | 0.842 | 0.986 | 0.947 |
| MNS | s | 0.976 | 1.000 | 0.996 | 0.998 | 0.988 |
| MNS | S | 0.993 | 0.998 | 0.998 | 0.993 | 0.995 |
| P1PK | P1 | 0.000 | 1.000 | NA | 0.774 | 0.500 |
| Rh | c | 0.000 | 1.000 | NA | 0.822 | 0.500 |
| Rh | C | 0.999 | 0.293 | 0.450 | 0.997 | 0.646 |
| Rh | $C^w$ | 1.000 | 0.835 | 0.994 | 0.987 | 0.918 |
| Rh | D | 0.977 | 0.591 | 0.382 | 0.990 | 0.784 |
| Rh | e | 0.964 | 1.000 | 0.992 | 0.999 | 0.982 |
| Rh | E | 0.999 | 0.985 | 0.994 | 0.998 | 0.992 |

[a] O, $A_1$, and $A_2$ in this column refer to phenotype.

#### Supplementary Table 6. Accuracy metrics for the Danish models in the Danish train data set.

| Blood group/HPA system | Antigen[a] | Sensitivity | Specificity | Positive predictive value | Negative predictive value | Balanced accuracy |
|---|---|---|---|---|---|---|
| ABO | A | 0.999 | 0.998 | 0.998 | 0.998 | 0.998 |
| ABO | AB | 1.000 | 0.997 | 1.000 | 0.996 | 0.998 |
| ABO | B | 1.000 | 0.997 | 1.000 | 0.998 | 0.999 |
| ABO | O | 0.999 | 0.997 | 0.998 | 0.999 | 0.998 |
| ABO | $A_1$ | 0.976 | 0.991 | 0.972 | 0.992 | 0.983 |
| ABO | $A_2$ | 0.987 | 0.797 | 0.929 | 0.959 | 0.892 |
| Cartwright | $Yt^a$ | 1.000 | 1.000 | 1.000 | 1.000 | 1.000 |
| Cartwright | $Yt^b$ | 1.000 | 0.993 | 0.999 | 0.996 | 0.996 |
| Colton | $Co^a$ | 0.500 | 0.999 | 0.600 | 0.999 | 0.750 |
| Colton | $Co^b$ | 0.994 | 0.934 | 0.993 | 0.942 | 0.964 |
| Dombrock | $Do^a$ | 0.999 | 0.999 | 0.998 | 0.999 | 0.999 |
| Dombrock | $Do^b$ | 0.998 | 1.000 | 1.000 | 1.000 | 0.999 |
| Duffy | $Fy^a$ | 0.995 | 0.998 | 0.995 | 0.998 | 0.997 |
| Duffy | $Fy^b$ | 0.959 | 0.999 | 0.995 | 0.990 | 0.979 |
| HPA-1 | HPA-1a | 1.000 | 1.000 | 1.000 | 1.000 | 1.000 |
| HPA-1 | HPA-1b | 1.000 | 1.000 | 1.000 | 1.000 | 1.000 |
| Kell | K | 0.996 | 0.911 | 0.992 | 0.953 | 0.954 |
| Kell | k | 0.562 | 1.000 | 0.932 | 0.996 | 0.781 |
| Kell | $Kp^a$ | 0.999 | 0.929 | 0.998 | 0.951 | 0.964 |
| Kell | $Kp^b$ | 0.600 | 1.000 | 1.000 | 1.000 | 0.800 |
| Kidd | $Jk^a$ | 0.993 | 0.998 | 0.995 | 0.998 | 0.996 |
| Kidd | $Jk^b$ | 0.995 | 0.999 | 0.997 | 0.998 | 0.997 |
| Knops | $Kn^a$ | 1.000 | 1.000 | 1.000 | 1.000 | 1.000 |
| Knops | $Kn^b$ | 1.000 | 1.000 | 1.000 | 1.000 | 1.000 |
| Lewis | $Le^a$ | 0.995 | 0.978 | 0.995 | 0.978 | 0.987 |
| Lewis | $Le^b$ | 0.487 | 0.974 | 0.936 | 0.712 | 0.731 |
| Lutheran | $Lu^a$ | 0.996 | 0.933 | 0.994 | 0.957 | 0.965 |
| Lutheran | $Lu^b$ | 0.563 | 1.000 | 1.000 | 0.998 | 0.781 |
| MNS | M | 0.988 | 0.998 | 0.993 | 0.997 | 0.993 |
| MNS | N | 0.983 | 0.990 | 0.976 | 0.993 | 0.987 |
| MNS | S | 0.997 | 0.998 | 0.998 | 0.997 | 0.998 |
| MNS | s | 0.988 | 1.000 | 0.995 | 0.999 | 0.994 |
| P1PK | P1 | 0.928 | 0.994 | 0.980 | 0.979 | 0.961 |
| Rh | C | 0.994 | 0.999 | 0.998 | 0.997 | 0.997 |
| Rh | c | 0.995 | 0.999 | 0.995 | 0.999 | 0.997 |
| Rh | D | 0.994 | 0.999 | 0.997 | 0.998 | 0.996 |
| Rh | E | 0.999 | 0.991 | 0.996 | 0.997 | 0.995 |
| Rh | e | 0.972 | 1.000 | 0.995 | 0.999 | 0.986 |
| Rh | $C^w$ | 0.999 | 0.907 | 0.997 | 0.959 | 0.953 |
| Vel | Vel | 0.676 | 1.000 | 1.000 | 0.999 | 0.838 |

[a] O, $A_1$, and $A_2$ in this column refer to phenotype.

#### Supplementary Table 7. Accuracy metrics for the Danish models in the Danish full data set.

| Blood group/HPA system | Antigen[a] | Sensitivity | Specificity | Positive predictive value | Negative predictive value | Balanced accuracy |
|---|---|---|---|---|---|---|
| ABO | A | 0.999 | 0.998 | 0.999 | 0.999 | 0.999 |
| ABO | AB | 1.000 | 0.998 | 1.000 | 0.997 | 0.999 |
| ABO | B | 1.000 | 0.997 | 1.000 | 0.998 | 0.999 |
| ABO | O | 0.999 | 0.998 | 0.998 | 0.999 | 0.998 |
| ABO | $A_1$ | 0.977 | 0.991 | 0.974 | 0.992 | 0.984 |
| ABO | $A_2$ | 0.987 | 0.795 | 0.929 | 0.959 | 0.891 |
| Cartwright | $Yt^a$ | 1.000 | 1.000 | 0.966 | 1.000 | 1.000 |
| Cartwright | $Yt^b$ | 1.000 | 0.996 | 1.000 | 0.996 | 0.998 |
| Colton | $Co^a$ | 0.667 | 1.000 | 0.800 | 0.999 | 0.833 |
| Colton | $Co^b$ | 0.995 | 0.930 | 0.993 | 0.954 | 0.963 |
| Dombrock | $Do^a$ | 0.999 | 0.999 | 0.999 | 1.000 | 0.999 |
| Dombrock | $Do^b$ | 0.998 | 1.000 | 0.999 | 1.000 | 0.999 |
| Duffy | $Fy^a$ | 0.996 | 0.998 | 0.996 | 0.998 | 0.997 |
| Duffy | $Fy^b$ | 0.961 | 0.999 | 0.996 | 0.991 | 0.980 |
| HPA-1 | HPA-1a | 1.000 | 1.000 | 1.000 | 1.000 | 1.000 |
| HPA-1 | HPA-1b | 0.997 | 1.000 | 1.000 | 0.994 | 0.999 |
| Kell | K | 0.996 | 0.915 | 0.993 | 0.953 | 0.956 |
| Kell | k | 0.676 | 0.999 | 0.925 | 0.997 | 0.838 |
| Kell | $Kp^a$ | 0.999 | 0.924 | 0.998 | 0.956 | 0.961 |
| Kell | $Kp^b$ | 0.500 | 1.000 | 1.000 | 1.000 | 0.750 |
| Kidd | $Jk^a$ | 0.995 | 0.998 | 0.995 | 0.998 | 0.997 |
| Kidd | $Jk^b$ | 0.995 | 0.999 | 0.997 | 0.998 | 0.997 |
| Knops | $Kn^a$ | 0.500 | 1.000 | 1.000 | 0.999 | 0.750 |
| Knops | $Kn^b$ | 1.000 | 1.000 | 1.000 | 1.000 | 1.000 |
| Lewis | $Le^a$ | 0.996 | 0.982 | 0.996 | 0.981 | 0.989 |
| Lewis | $Le^b$ | 0.490 | 0.982 | 0.954 | 0.714 | 0.736 |
| Lutheran | $Lu^a$ | 0.997 | 0.932 | 0.994 | 0.964 | 0.964 |
| Lutheran | $Lu^b$ | 0.656 | 1.000 | 0.955 | 0.999 | 0.828 |
| MNS | M | 0.989 | 0.998 | 0.994 | 0.997 | 0.993 |
| MNS | N | 0.982 | 0.991 | 0.976 | 0.993 | 0.987 |
| MNS | S | 0.997 | 0.997 | 0.997 | 0.997 | 0.997 |
| MNS | s | 0.986 | 1.000 | 0.995 | 0.999 | 0.993 |
| P1PK | P1 | 0.934 | 0.995 | 0.983 | 0.981 | 0.965 |
| Rh | C | 0.995 | 0.999 | 0.999 | 0.997 | 0.997 |
| Rh | c | 0.996 | 0.999 | 0.995 | 0.999 | 0.997 |
| Rh | D | 0.994 | 0.999 | 0.997 | 0.998 | 0.997 |
| Rh | E | 0.999 | 0.991 | 0.997 | 0.997 | 0.995 |
| Rh | e | 0.969 | 1.000 | 0.993 | 0.999 | 0.984 |
| Rh | $C^w$ | 0.999 | 0.908 | 0.997 | 0.965 | 0.953 |
| Vel | Vel | 0.622 | 1.000 | 0.920 | 0.999 | 0.811 |

[a] O, $A_1$, and $A_2$ in this column refer to phenotype.

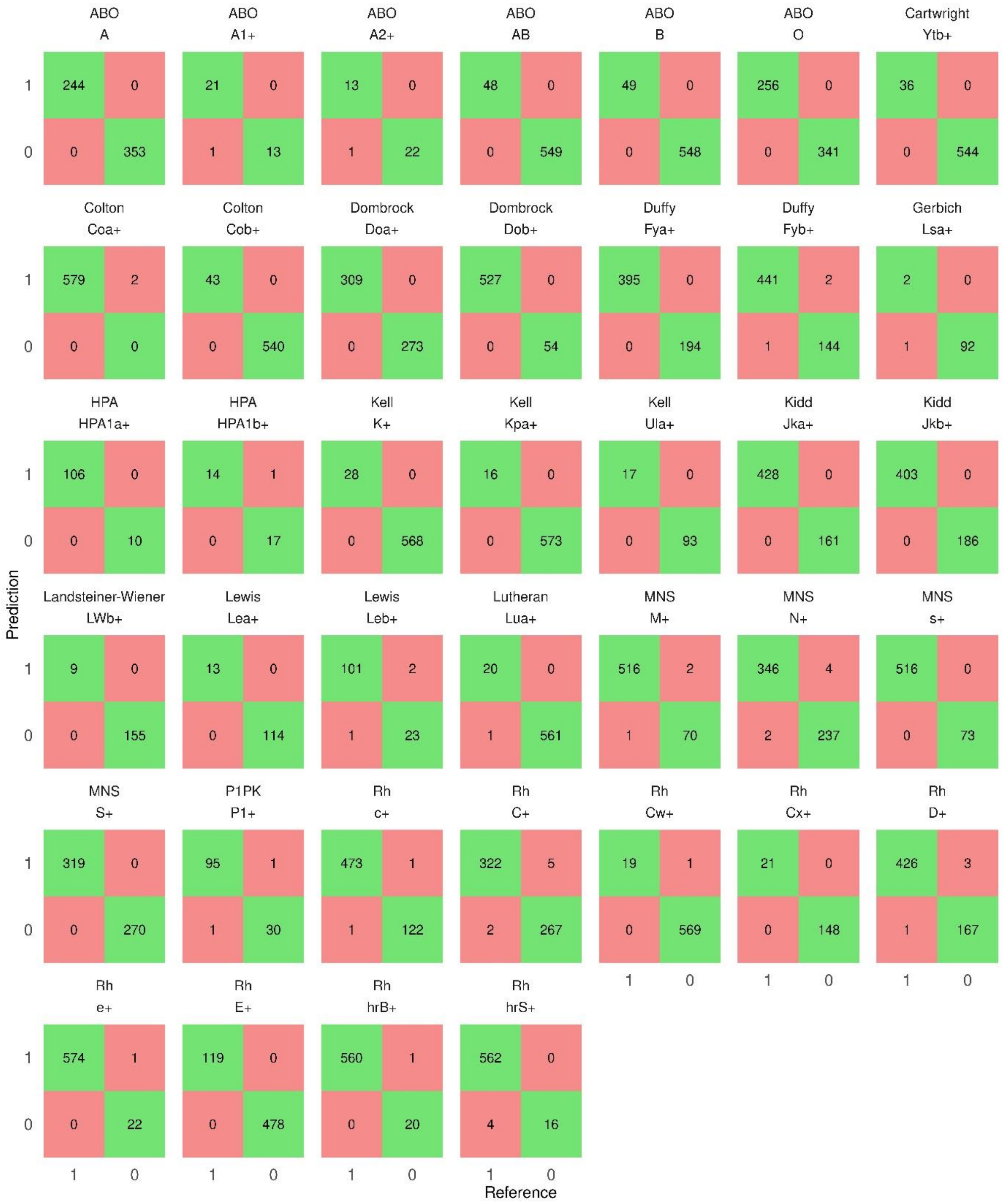

#### Supplementary Figure 1. Confusion matrices for the Finnish models in the Finnish train data set.

Confusion matrices for the Finnish antigen classification models in the Finnish train data set are presented in alphabetical order of the blood group systems. The RBC antigen/phenotype and HPA-1 typing results are on the x-axis and the model predictions on the y-axis. The antigen-negative samples are denoted by 0 and the antigen-positive samples by 1 on both axes. The numbers of true positive and true negative samples are depicted in the green boxes and the numbers of false positive and false negative samples in the red boxes.

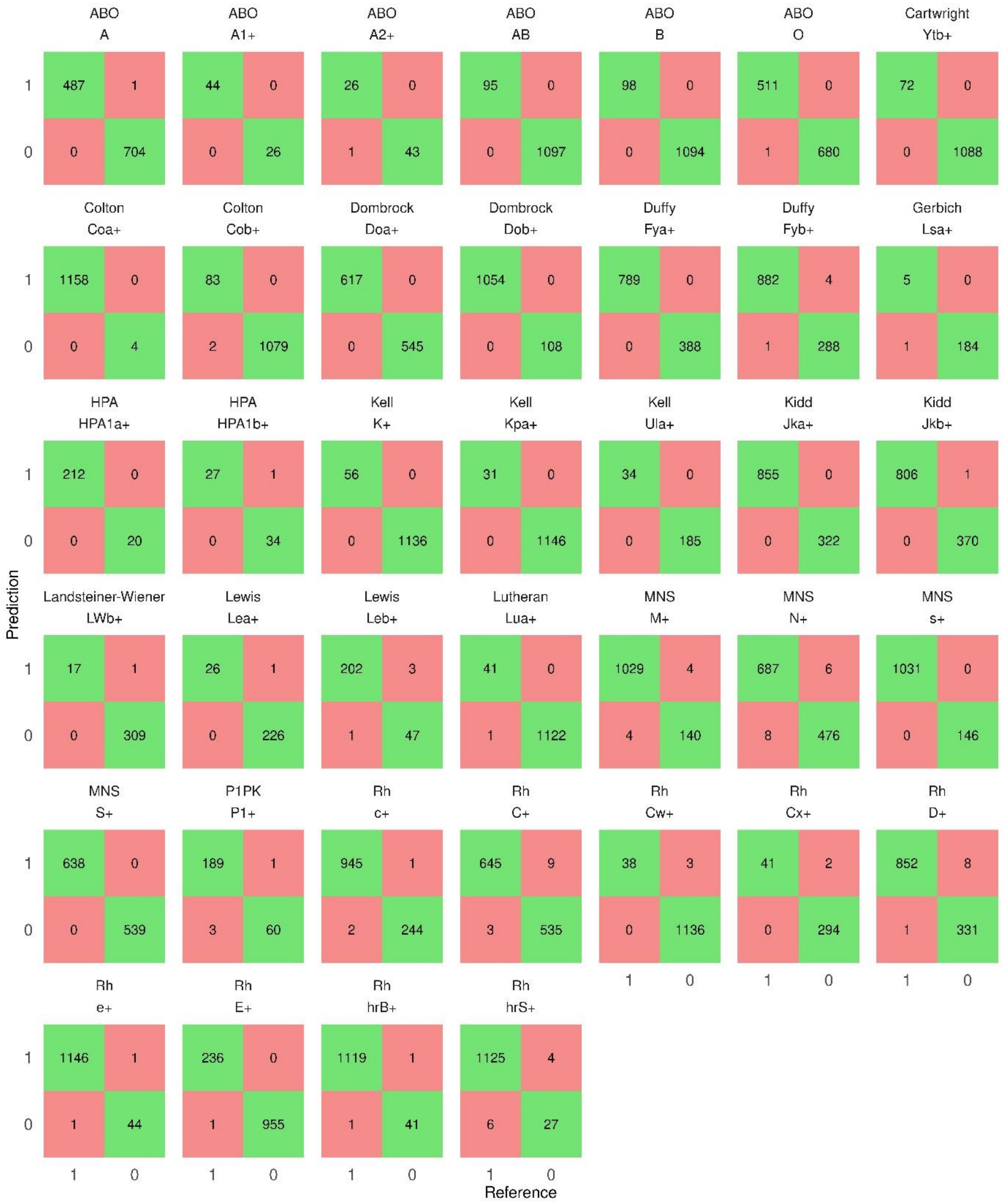

#### Supplementary Figure 2. Confusion matrices for the Finnish models in the Finnish full data set

Confusion matrices for the Finnish antigen classification models in the Finnish full data set are presented in alphabetical order of the blood group systems. The RBC antigen/phenotype and HPA-1 typing results are on the x-axis and the model predictions on the y-axis. The antigen-negative samples are denoted by 0 and the antigen-positive samples by 1 on both axes. The numbers of true positive and true negative samples are depicted in the green boxes and the numbers of false positive and false negative samples in the red boxes.

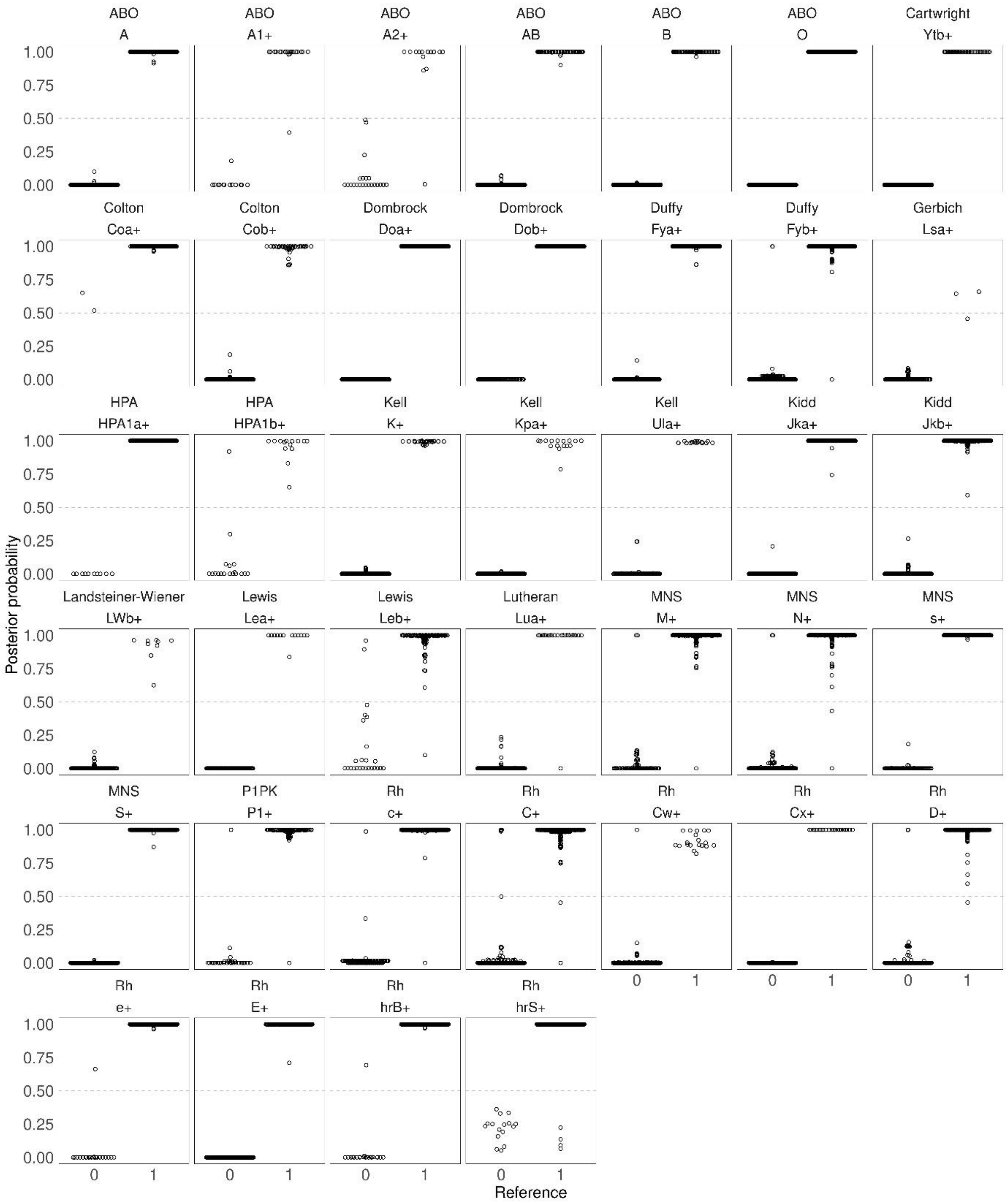

#### Supplementary Figure 3. Posterior probability boxplots for the Finnish models in the Finnish train data set

Posterior probability boxplots for the Finnish antigen classification models in the Finnish train data set are presented in alphabetical order of the blood group systems. The RBC antigen/phenotype and HPA-1 typing results are on the x-axis and the antigen-negative samples are denoted by 0 and the antigen-positive samples by 1. The posterior probabilities for samples range from 0 to 1 and are presented on the y-axis. Samples are depicted as open circles.

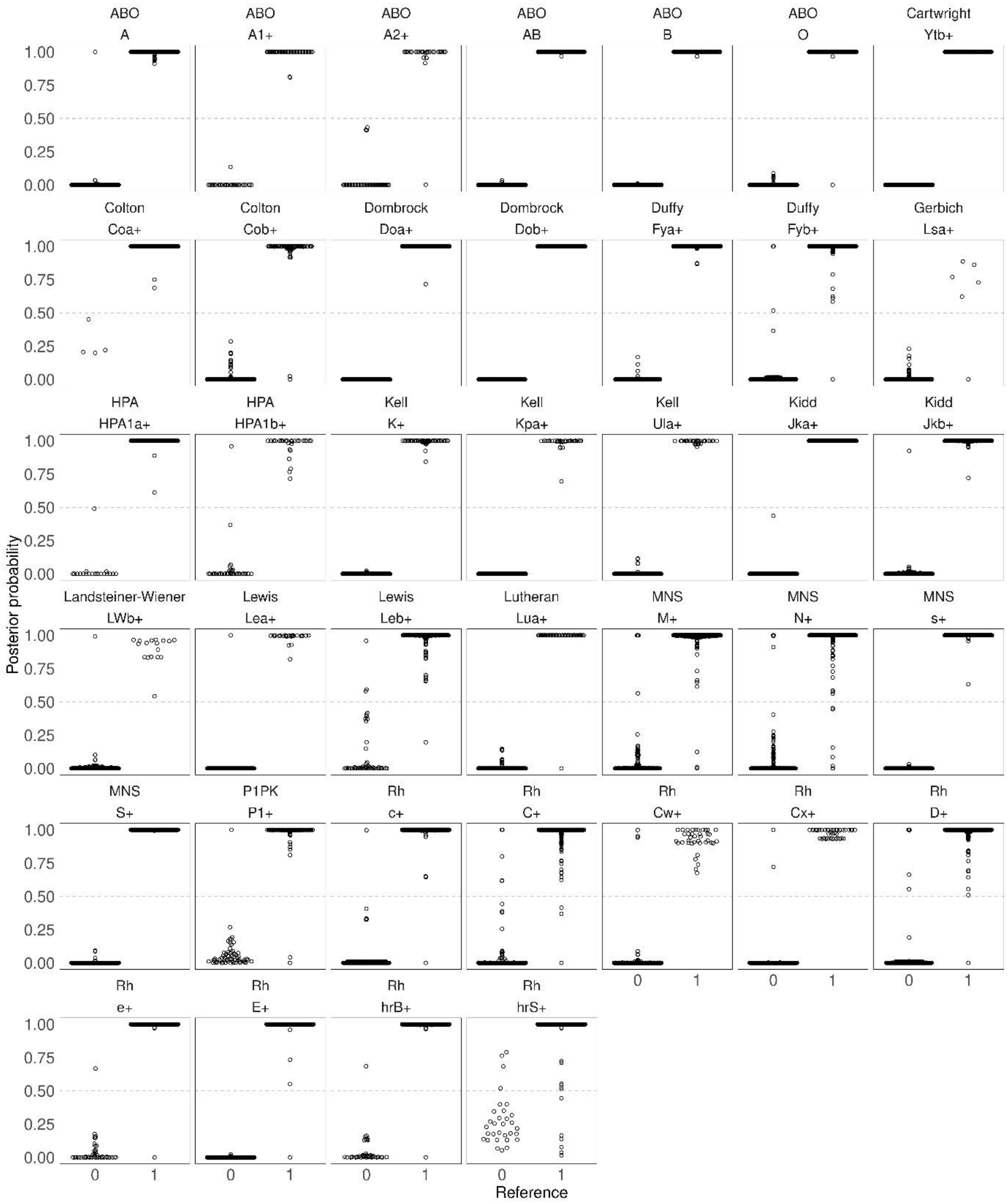

#### Supplementary Figure 4. Posterior probability boxplots for the Finnish models in the Finnish full data set.

Posterior probability boxplots for the Finnish antigen classification models in the Finnish full data set are presented in alphabetical order of the blood group systems. The RBC antigen/phenotype and HPA-1 typing results are on the x-axis and the antigen-negative samples are denoted by 0 and the antigen-positive samples by 1. The posterior probabilities for samples range from 0 to 1 and are presented on the y-axis. Samples are depicted as open circles.

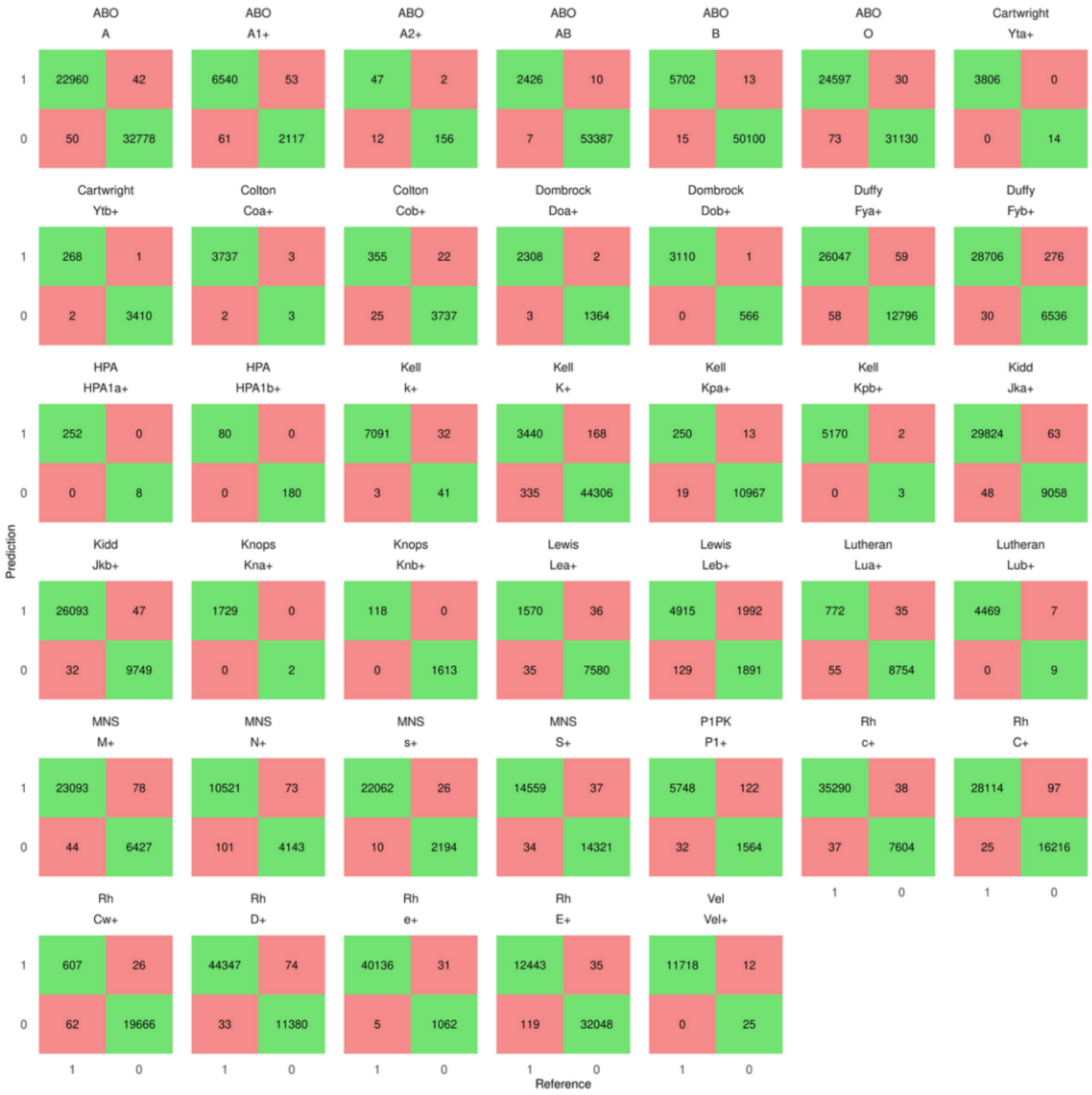

#### Supplementary Figure 5. Confusion matrices for the Danish models in the Danish train data set

Confusion matrices for the Danish antigen classification models in the Danish train data set are presented in alphabetical order of the blood group systems. The RBC antigen/phenotype and HPA-1 typing results are on the x-axis and the model predictions on the y-axis. The antigen-negative samples are denoted by 0 and the antigen-positive samples by 1 on both axes. The numbers of true positive and true negative samples are depicted in the green boxes and the numbers of false positive and false negative samples in the red boxes.

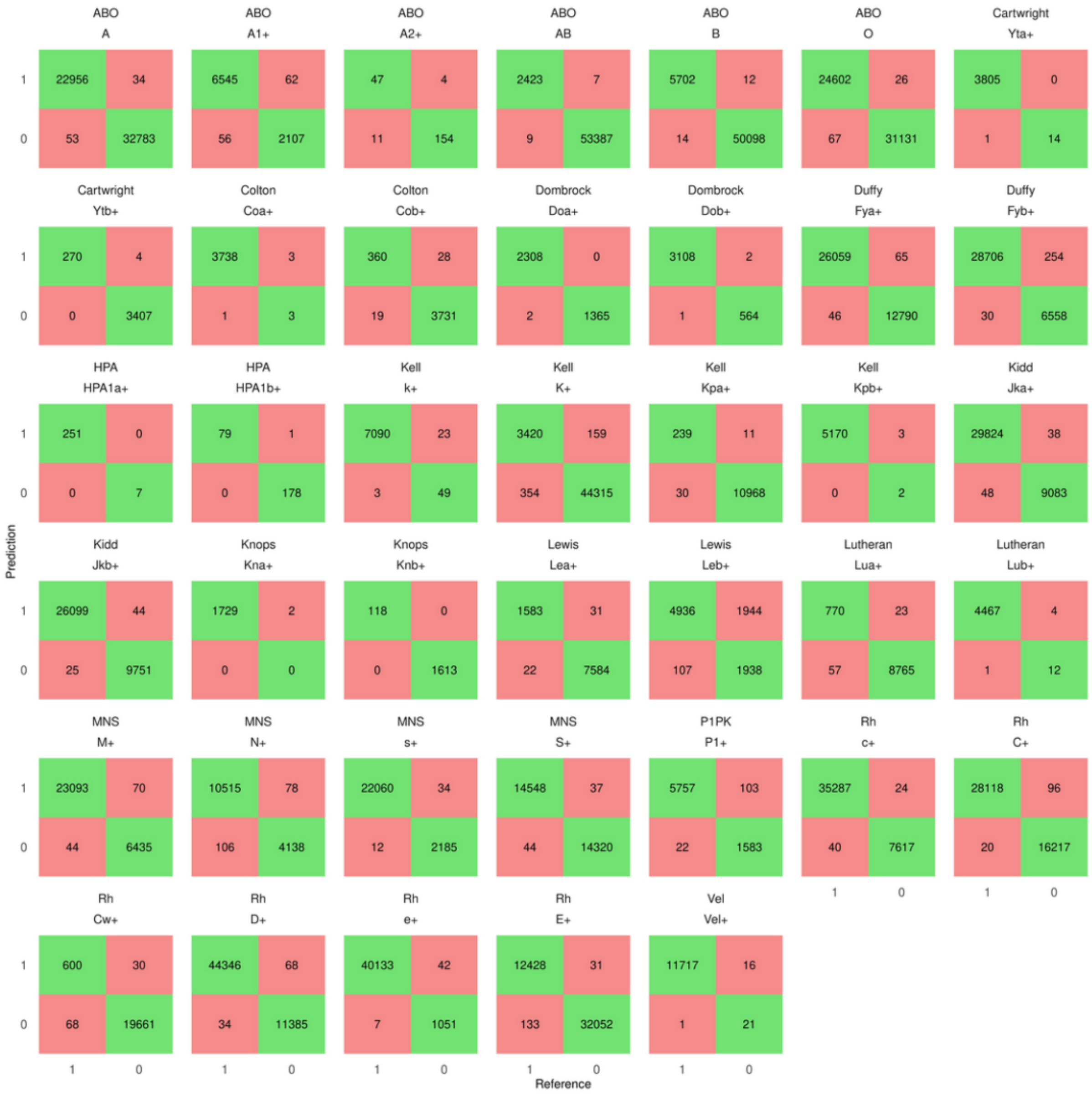

#### Supplementary Figure 6. Confusion matrices for the Danish models in the Danish test data set

Confusion matrices for the Danish antigen classification models in the Danish test data set are presented in alphabetical order of the blood group systems. The RBC antigen/phenotype and HPA-1 typing results are on the x-axis and the model predictions on the y-axis. The antigen-negative samples are denoted by 0 and the antigen-positive samples by 1 on both axes. The numbers of true positive and true negative samples are depicted in the green boxes and the numbers of false positive and false negative samples in the red boxes.

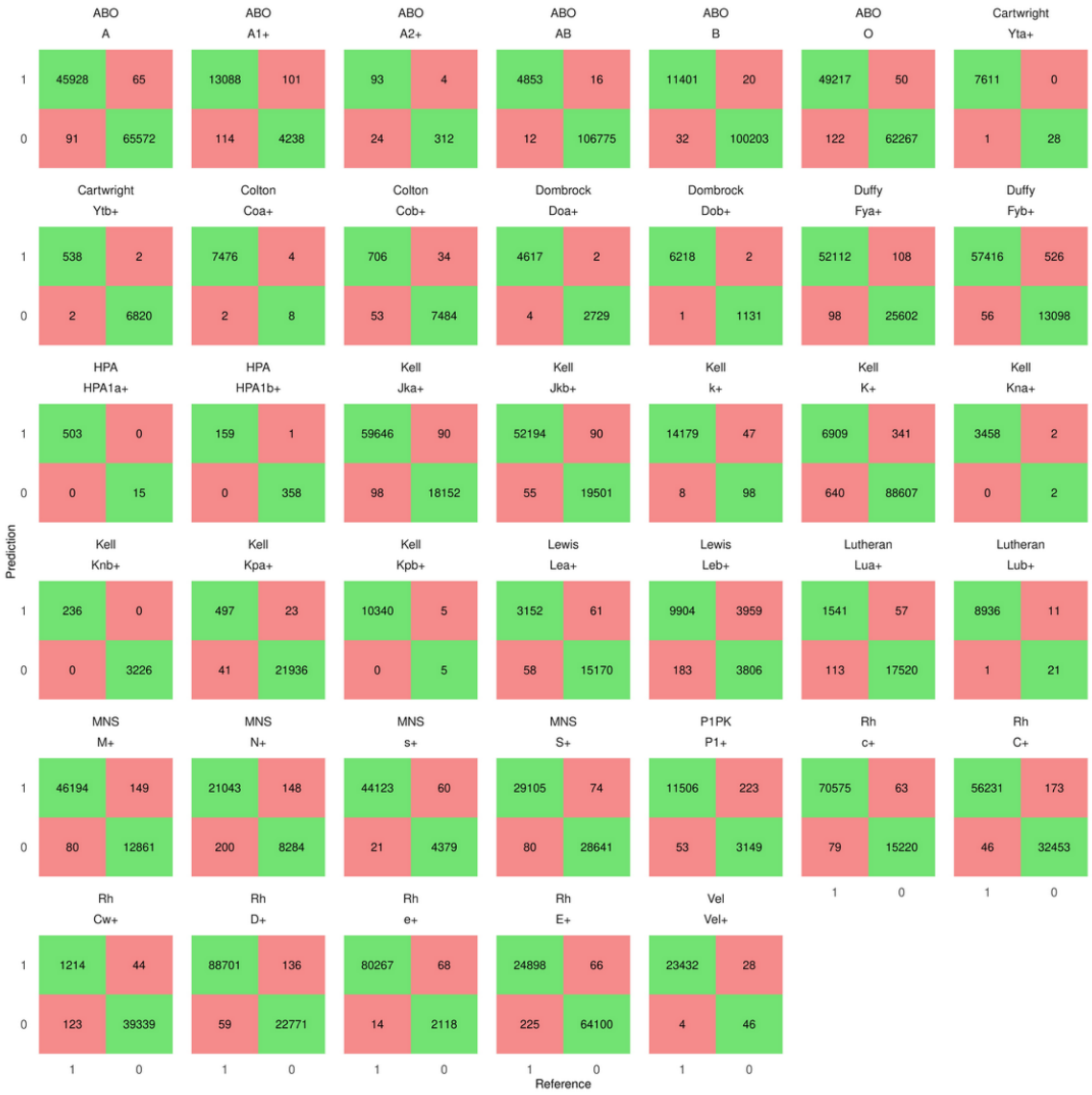

#### Supplementary Figure 7. Confusion matrices for the Danish models in the Danish full data set

Confusion matrices for the Danish antigen classification models in the Danish full data set are presented in alphabetical order of the blood group systems. The RBC antigen/phenotype and HPA-1 typing results are on the x-axis and the model predictions on the y-axis. The antigen-negative samples are denoted by 0 and the antigen-positive samples by 1 on both axes. The numbers of true positive and true negative samples are depicted in the green boxes and the numbers of false positive and false negative samples in the red boxes.
